# *In Vivo* Assessment of the Safety of Standard Fractionation Temporally Feathered Radiation Therapy (TFRT) for Head and Neck Squamous Cell Carcinoma: An R-IDEAL Stage 1/2a First-in-Humans/Feasibility Demonstration of New Technology Implementation

**DOI:** 10.1101/2021.01.05.21249232

**Authors:** Shireen Parsai, Richard Lei J. Qiu, Peng Qi, Geoffrey Sedor, Clifton D. Fuller, Eric Murray, David Majkszak, Nicole Dorio, Shlomo Koyfman, Neil Woody, Nikhil Joshi, Jacob G. Scott

**Author notes:** Co-corresponding Authors: Jacob G. Scott, 10201 Carnegie Ave, Cleveland, OH 44195, Nikhil Joshi, MD, 500 S Paulina St Suite 013, Chicago, IL 60612.

## Abstract

**Introduction:** Prior *in silico* simulations of studies of Temporally Feathered Radiation Therapy (TFRT) have demonstrated potential reduction in normal tissue toxicity. This R-IDEAL Stage 1/2A study seeks to demonstrate the first-in-human implementation of TFRT in treating patients with head and neck squamous cell carcinoma (HNSCC).

**Materials and Methods:** Patients with HNSCC treated with definitive radiation therapy were eligible (70 Gy in 35 fractions) were eligible. The primary endpoint was feasibility of TFRT planning as defined by radiation start within 15 days of CT simulation. Secondary endpoints included estimates of acute grade 3-5 toxicity.

**Results:** The study met its accrual goal of 5 patients. TFRT plans were generated in four of the five patients within 15 business days of CT simulation, therefore meeting the primary endpoint. One patient was not treated with TFRT at physician’s discretion, though the TFRT plan had been generated within sufficient time from the CT simulation. For patients who received TFRT, the median time from CT simulation to radiation start was 10 business days (range 8-15). The average time required for radiation planning was 6 days. In all patients receiving TFRT, each subplan and every daily fraction was delivered in the correct sequence without error. The OARs feathered included: oral cavity, each submandibular gland, each parotid gland, supraglottis, and posterior pharyngeal wall (OAR pharynx). Prescription dose PTV coverage (>95%) was ensured in each TFRT subplan and the composite TFRT plan. One of five patients developed an acute grade 3 toxicity.

**Conclusions:** This study demonstrates the first-in-human implementation of TFRT (R-IDEAL Stage 1), proving its feasibility in the modern clinical workflow. Additionally, assessments of acute toxicities and dosimetric comparisons to a standard radiotherapy plan were described (R-IDEAL Stage 2a).

**Highlights:**

- This prospective study is the first-in-human application for Temporally Feathered Radiation Therapy (TFRT).
- In theory, TFRT may reduce radiation-induced toxicities by optimizing the time through which radiation is delivered and consequently improve normal tissue recovery.
- In this study, patients with head and neck squamous cell carcinoma were treated with TFRT.
- The primary endpoint of technical feasibility was met when patients were successfully treated with TFRT techniques without introducing delays in radiation commencement.

## Introduction

Radiation plays an important role in the definitive treatment of various head and neck cancers.(1-3) Conventional techniques of radiotherapy result in high rates of late effects, such as dysphagia, radionecrosis, and xerostomia. The physical dose distributions over the target volume and nearby surrounding structures has improved with the advancement of radiation therapy techniques and specifically the adoption of intensity-modulated radiotherapy (IMRT) coupled with image guidance.(4) This has led to decreased toxicity rates, especially xerostomia.(5) In a recent study of patients receiving modern techniques of IMRT with or without chemotherapy for HPV-related oropharynx cancer, 42% of patients experienced grade 3 or higher acute toxicity with the most common being dysphagia (24%), mucositis (17%), and dermatitis (8%).(6) Of the patients followed greater than one year without local failure, 24% experienced grade 2 xerostomia. Late high grade toxicity remained low in this study. Quality of life also mirrors the decline in physical functioning. Though most patients recover global quality of life by 12 months, deterioration in physical functioning, fatigue, xerostomia, and sticky saliva persist beyond 12 months in head and neck cancer survivors.(7)

Temporally Feathered Radiation Therapy (TFRT) has been proposed as a technique to reduce toxicity in patients undergoing radiation therapy to the head and neck.(8) As IMRT previously optimized the physical distribution of radiation dose, TFRT seeks to optimize the time through which radiation therapy is delivered and take advantage of the non-linear recovery of normal tissues. With the TFRT technique, a higher fractional dose is delivered once weekly to an organ at risk (OAR) and a lower fractional radiation dose is delivered for the remaining 4 fractions. *In silico* simulations have demonstrated opportunities for greater normal tissue recovery by increasing the time between days the OAR receives a higher fractional radiation dose.(8) In this study of TFRT planning, 5 OARs are feathered through time in this fashion with one OAR being deprioritized and receiving higher fractional dose per day. In clinical practice, this would require 5 separate radiation subplans, each deprioritizing a different OAR, and each delivered once per day of the week (Figure 1). The TFRT plan is considered to be the composite of these 5 subplans. This increases the complexity of planning and treatment delivery.

**Figure 1.**
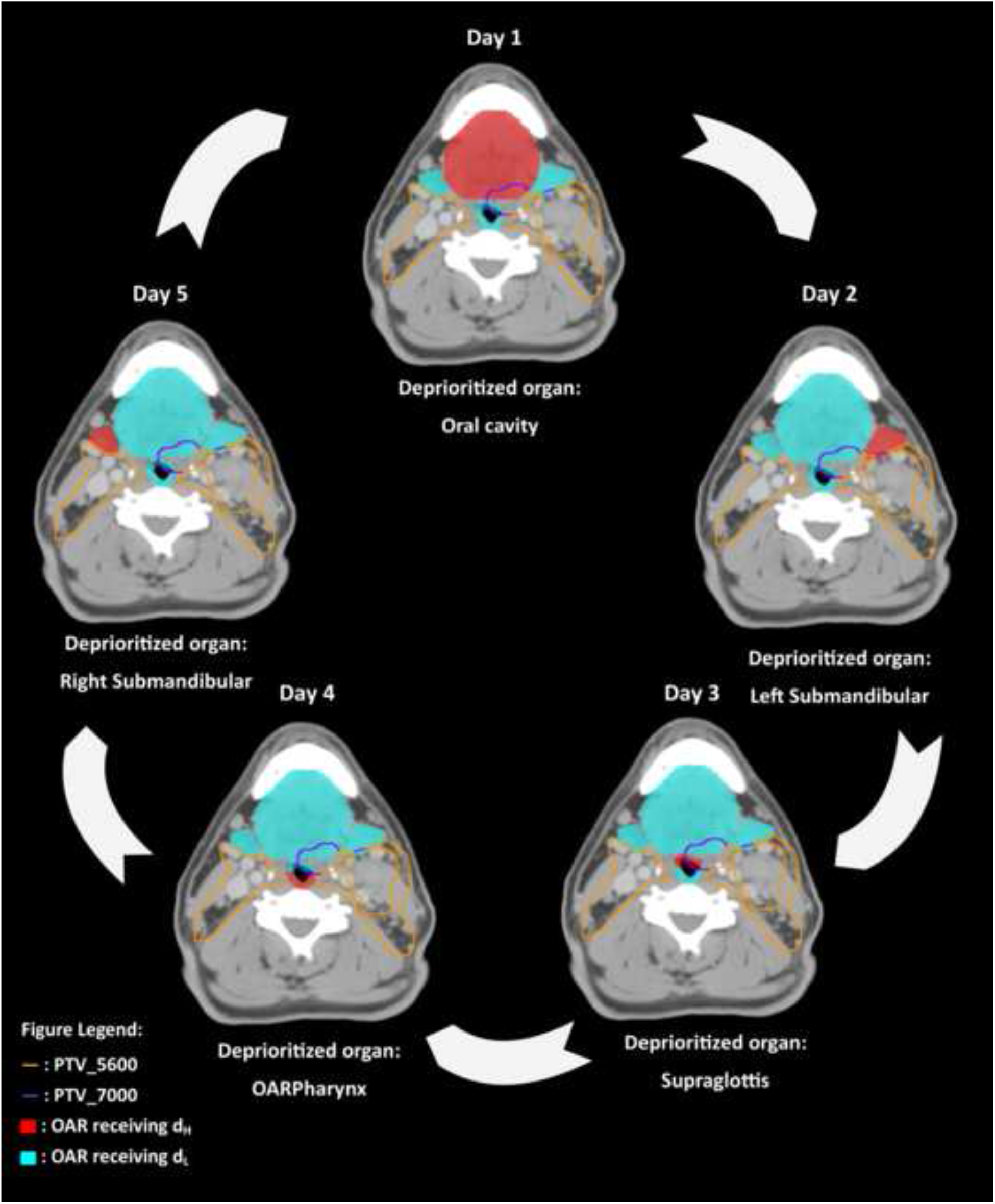
Schematic representation of Temporally Feathered Radiation Therapy. Five individual radiation plans, aka subplans, are created corresponding to each day of the week. In each subplan, one OAR is deprioritized such that it receives a higher fractional dose, d_H_, shown in red. The OARs surrounding the high dose PTV receiving a lower fractional dose, d_L_, are shown in blue. The high dose PTV, receiving 70 Gy, is outlined in purple, whereas the elective volume receiving 56 Gy is outlined in orange. A composite TFRT plan is created by the dose accumulation of the five subplans.

In addition to undertaking this pilot of standard fractionation radiotherapy using the TFRT approach, we used the previously described R-IDEAL conceptual framework for technology development in radiotherapy.(9) As per R-IDEAL, we sought to implement a structured evaluation of a novel radiotherapy technology/approach, namely, TFRT, in a programmatic prospective implementation series. Consistent with R-IDEAL methodology, this implementation assessment is consistent with R-IDEAL Stage 1 (“Idea”, i.e. first-in-human implementation as structured case reportage) and R-IDEAL Stage 2a (i.e. “Development”, i.e. demonstration of technical feasibility “in which additional modifications are made to further optimize work-flow and technology for innovative treatment delivery”, as uninterrupted cases series). The use of the R-IDEAL framework allows iterative technology development to be systematically reported serially as continuous effort, rather than *ad hoc* presentations of retrospective single-institution experiences, and is coherent with the IDEAL surgical technology development framework which has seen increasing adoption.(10)

In a previous manuscript (8) we demonstrated the theoretical potential for gain in normal tissue complication probability (NTCP) with TFRT (Stage 0 of R-IDEAL). The specific aim of the current study is to demonstrate the first-in-human implementation of temporally feathered radiotherapy for standard fractionation radiotherapy in head and neck cancer (R-IDEAL Stage 1). Additionally, we aim to evaluate and report the technical feasibility of TFRT planning and delivery within standard current clinical workflow and commercial treatment planning systems, with the secondary endpoint of assessment of acute radiotherapy attributable toxicity, reported as a sequential case series (R-IDEAL Stage 2a). A complete comparative dosimetric analysis is also provided between the TFRT plans generated and conventional non-TFRT plans.

## Materials and Methods

### Participants

We undertook an Institutional Review Board (IRB #18-1248) approved, structured as an R-IDEAL Stage 1/2a technical feasibility assessment. Eligible patients had histologically confirmed non-metastatic squamous cell carcinoma of the oropharynx, nasopharynx, larynx, and hypopharynx planned to be treated with definitive radiation therapy to 70 Gy in 35 fractions. Concurrent chemotherapy was allowed. Karnofsky Performance Status (KPS) of 80 or greater was required. Patients were excluded on the basis of receiving any other investigational agents, prior oncologic surgery, or prior head and neck radiation therapy. All patients were provided written informed consent. The protocol schema is included in the supplementary material (Appendix A).

### Procedures

The gross tumor volumes (GTV), clinical target volumes (CTV), planning target volumes (PTV) and all normal structures were delineated by the radiation oncologist as previously described.(6,11) 70 Gy in 35 fractions was delivered to the primary tumor volume. Nodal groups at risk for harboring micrometastatic disease received 56 Gy in 35 fractions. An intermediate dose level of 63 Gy in 35 fractions was allowed. A simultaneous integrated boost technique was used. Pinnacle^3^ treatment planning software (version 9.10, Philips Healthcare, Fitchburg, WI) was used for all planning.

The primary endpoint of the study was the feasibility of TFRT planning and delivery, defined as commencing TFRT within 15 days of simulation. Following CT simulation TFRT plans were generated, while in tandem a non-TFRT IMRT plan was generated as per regular institutional practice by a separate group of radiation planners. If the TFRT plan was not completed in time, the patient was treated with the non-TFRT IMRT plan to avoid delays in patient treatment start. Details of TFRT plan generation are previously described.(11) Before TFRT planning, the treating physician and a physicist identified five OARs to be feathered based on proximity to the target. These OARs may have included but were not limited to the oral cavity, each submandibular gland, each parotid gland, OAR pharynx, supraglottis, larynx, and esophagus. The compliance criteria and suggested dosimetric constraints can be found in the supplementary material (Appendix B). Quality assurance was performed on each TFRT subplan. An additional time out process was carried out by the radiation therapists prior to the delivery of daily TFRT plans (Appendix C). Secondary endpoints included estimates of acute grade 3-5 toxicity (within 3 months after radiation), as graded per CTCAE criteria version 4. Lastly, dosimetric data was also gathered as part of exploratory endpoints.

### Endpoint Definitions and Data Collection

The This R-IDEAL Stage 1/2a study predefined technical threshold to meet feasibility was set as completion and delivery of TFRT plans in three patients within a standard radiation planning interval, with treatment completion without technical plan modification or physician-reported adverse events. A standard radiation planning interval was defined additionally such that no greater than 15 business days were allowed to elapse between CT simulation date and treatment start date was deemed clinically acceptable. Each TFRT subplan was required to be delivered in the correct, pre-specified sequence, without technical modification, machine error per standard. Descriptive statistics were used to report treatment-related toxicity. Dosimetric analyses were conducted using descriptive statistics. Toxicity Data was gathered in a RedCap database.

## Results

The study met its accrual goal of 5 patients. Table 1 shows the patient and tumor characteristics. All patients enrolled had a diagnosis of p16+ oropharyngeal squamous cell carcinoma and received 70 Gy in 35 fractions to the primary tumor and 56 Gy in 35 fractions to bilateral neck to the nodal levels at risk for micrometastatic disease; one patient was treated with an additional intermediate dose of 63 Gy in 35 fractions. TFRT plans were generated in four of the five patients within 15 business days of CT simulation, therefore meeting the primary endpoint of technical feasibility. The median time from CT simulation date to radiation start date was 10 business days (range 8-15) for patients who received TFRT. One patient was opted to not be treated by TFRT at the physician’s discretion. In this case, the PTV_7000 volume was the smallest (32 cc) and therefore the surrounding OARs were already receiving favorable doses. It is important to note that the TFRT plan had been generated within sufficient time from the CT simulation for this patient and met all compliance criteria previously referenced in Appendix B. The average planning time by the dosimetrists and physicists was 6 days. In all patients receiving TFRT, each subplan and every daily fraction was delivered in the correct sequence without error. Three patients required adjustment of the treatment schedule in the record and verify system due to holidays. One of four patients who received TFRT underwent an adaptive replan at the physician’s discretion due to tumor shrinkage and anatomical changes. The replan also implemented TFRT technique. The time from simulation to the start of the adaptive replan was 6 business days, the time required for planning was 4 days.

**Table 1.** Patient and tumor characteristics.

|  |  |
| --- | --- |
| Median age | 66.8 (56.4-69.2) |
| Number of men | 5 |
| KPS |  |
| 90 | 4 |
| 80 | 1 |
| Concurrent chemotherapy | 4 |
| Median PTV volume | 97.5 cc (range: 32.1-138.8) |
| Subsite |  |
| Base of Tongue | 4 |
| Tonsil | 1 |
| T stage* |  |
| T1 | 1 |
| T2 | 3 |
| T3 | 0 |
| T4 | 1 |
| N stage* |  |
| N0 | 1 |
| N1 | 3 |
| N2 | 1 |
| N3 | 0 |

\*TNM staging is per AJCC 8<sup>th</sup> edition. No patients with distant metastatic disease (M1) were included. All patients enrolled had HPV-related oropharyngeal squamous cell carcinoma.

### Treatment Delivery

OARs feathered included: oral cavity, each submandibular gland, each parotid gland, supraglottis, and posterior pharyngeal wall (OAR pharynx). The feathered OARs were chosen based on proximity to the PTV_7000 by the physician. The oral cavity, at least one submandibular gland, supraglottis, and OAR pharynx were feathered in every patient. Other OARs that were permitted to be feathered per protocol included the larynx and esophagus. The average magnitude of fluctuation between d_H_ and d_L_ was 17 cGy for the oral cavity (range 4-23), 34 cGy for the submandibular glands (range 24-46), 19 cGy for the supraglottis (range 4-27), 15 cGy for the OAR pharynx (range 7-29), and 24 cGy for the parotid glands (range 21-30). Figure 2 also demonstrates the daily fluctuation between d_H_ and d_L_ for every feathered OAR for each patient receiving TFRT. Of the 20 feathered organs (in 4 patients), 7 organs received higher total composite doses in the TFRT plans as compared to the non-TFRT IMRT plans. The differences between the composite TFRT plans and the non-TFRT IMRT plans are shown in Table 2. All additional dosimetric data is reported in the supplementary material (Appendix D).

**Table 2.** Difference in dose delivered to organs at risk between the TFRT composite plan and non-TFRT IMRT plan. The cells highlighted in grey indicate the feathered organs per plan. Negative values indicate less dose in the composite TFRT plan.

|  | Patient 1,<br>difference in<br>dose (cGy) | Patient 2,<br>difference in<br>dose (cGy) | Patient 3,<br>difference in<br>dose (cGy) | Patient 4,<br>difference in<br>dose (cGy) | Average<br>difference in dose<br>across all patients<br>(cGy) |
| --- | --- | --- | --- | --- | --- |
| Oral Cavity | -162 | -142 | -561 | 22 | -211 |
| Right Parotid | -87 | -251 | -153 | -502 | -248 |
| Left Parotid | 9 | -6 | -113 | 2 | -27 |
| Right<br>Submandibular | -328 | 24 | -177 | -494 | -199 |
| Left<br>Submandibular | 74 | -285 | 24 | 79 | -27 |
| OAR Pharynx | 293 | 41 | -162 | -204 | -8 |
| Esophagus | 47 | -291 | -435 | -61 | -185 |
| Supraglottis | -247 | 25 | 3 | -164 | -40 |
| Larynx | 261 | -83 | 33 | 133 | 86 |

**Figure 2.**
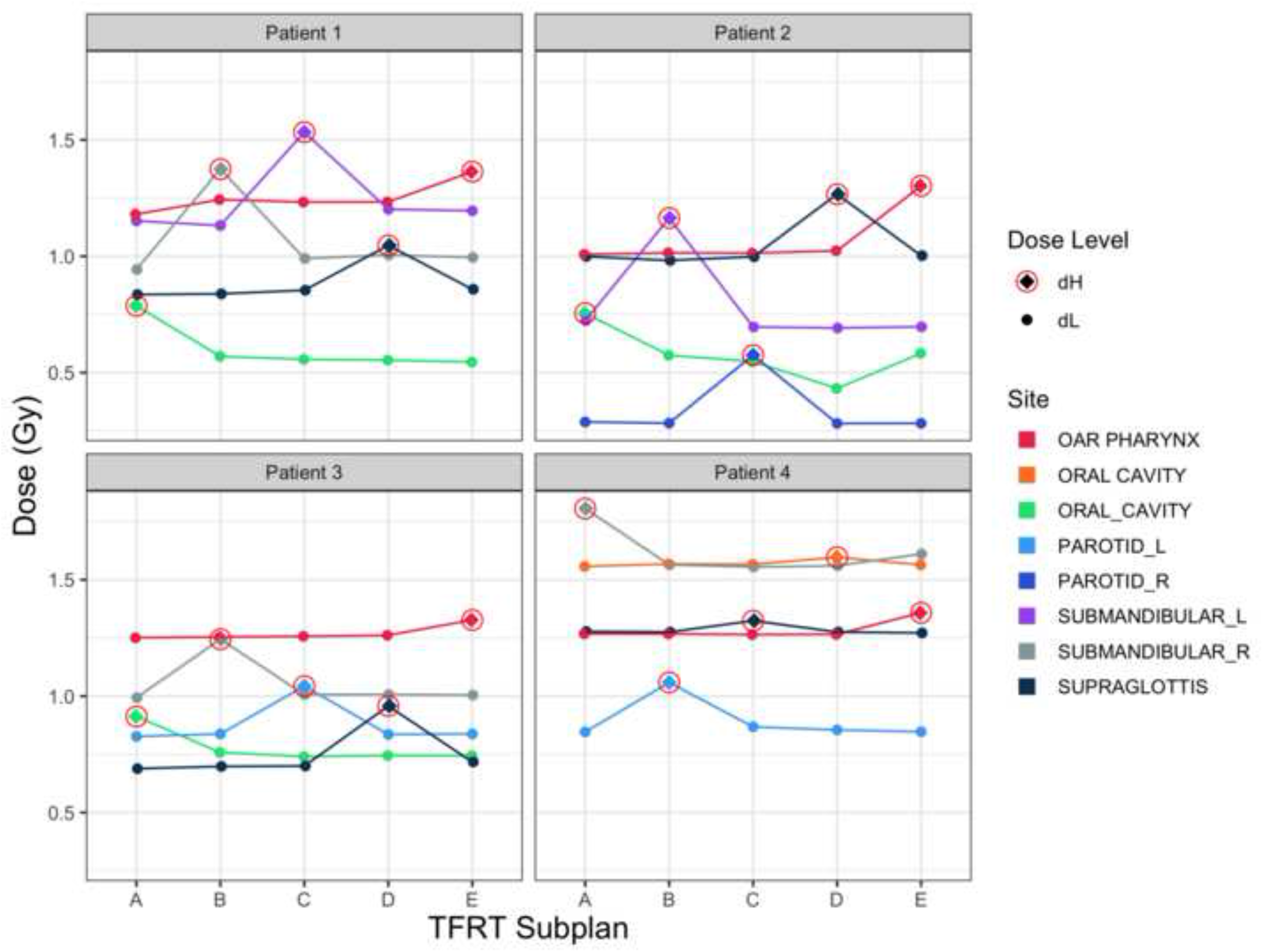
Fluctuation in daily dose delivered to the feathered OARs. The higher fractional dose delivered once weekly, d_H_, is highlighted with a red circle. The lower fractional doses, d_L_, are kept generally constant the remaining 4 fractions weekly. The average magnitude of fluctuation between d_H_ and d_L_ was 17 cGy for the oral cavity, 34 cGy for the submandibular glands, 19 cGy for the supraglottis, 15 cGy for the OAR pharynx, and 24 cGy for the parotid glands.

The same compliance criteria and planning objectives (Appendix B) were applied for both TFRT plans and non-TFRT IMRT plans. As is standard in non-TFRT IMRT plans, prescription dose PTV coverage (>95%) was ensured in each TFRT subplan and the composite TFRT plan. As a measure of homogeneity, D1cc 577 Gy was achieved for PTV_7000 in all plans. Conformality index was calculated for PTV_7000 and PTV_5600 as the prescription isodose volume divided by the planning target volume. The conformality index for PTV_7000 and PTV_5600 was 52.1 for all non-TFRT plans and composite TFRT plans, with one exception.

### Toxicity Assessment

One of five patients developed an acute grade 3 toxicity. This occurred in a patient with a large T4N1 oropharyngeal tumor (130 cc), with baseline dysphagia at presentation. Following treatment, the patient had worsened grade 3 dysphagia and vellopalatine insufficiency which required placement of a temporary feeding tube. No other high grade toxicities were observed. All acute toxicities can be referenced in the Table 3.

**Table 3.** Maximum toxicities recorded for patients by CTCAE v4 criteria.

| Maximum acute side effects graded by CTCAE v4 |  |  |  |  |  |
| --- | --- | --- | --- | --- | --- |
| Side effect | grade 0 | grade 1 | grade 2 | grade 3 | grade 4 |
| Dysphagia | 0 | 3 | 0 | 1* | 0 |
| Mucositis | 0 | 2 | 3 | 0 | 0 |
| Dermatitis | 0 | 3 | 2 | 0 | 0 |
| Xerostomia | 0 | 2 | 3 | 0 | 0 |
| Fatigue | 0 | 3 | 2 | 0 | 0 |
| Nausea | 2 | 3 | 0 | 0 | 0 |
| Vomiting | 4 | 0 | 0 | 0 | 0 |
| Pain | 0 | 2 | 3 | 0 | 0 |
| Taste | 0 | 2 | 3 | 0 | 0 |
| Weight Loss | 0 | 2 | 3 | 0 | 0 |

\*The only high grade toxicity was grade 3 dysphagia occurred in a patient who had a large T4N1 oropharyngeal tumor (130 cc), with baseline dysphagia at presentation.

## Discussion

This study represents the first-in-human delivery of Temporally Feathered Radiation Therapy in head and neck cancer (R-IDEAL Stage 1). We prospectively demonstrated the feasibility of treatment delivery in the current clinical workflow, thereby achieving the primary endpoint of the study. All TFRT subplans were administered in correct succession by the radiation therapists, including in events which required treatment calendar modifications. Planning target volume coverage (>95%) with prescription dose was achieved in every TFRT subplan and composite TFRT plan. No excess toxicity was observed as compared to prior institutional analysis.(6) This study lays the framework for further clinical research exploring TFRT.

Dosimetric comparisons between the TFRT plan and the non-TFRT IMRT plan were made (R-IDEAL Stage 2a). This was done by using the composite TFRT plan, created by arithmetic dose accumulation of the individual subplans, as a representation of the dose delivered by the TFRT technique. It was previously postulated that TFRT planning would result in greater total dose delivered to the feathered organs.(8) Additionally, it was postulated that despite greater radiation doses delivered to the feathered OARs, the normal tissue toxicity induced by radiation might be less because of improved normal tissue recovery from the temporal fluctuation of dose to the organs at risk.(8) This was simulated using a dynamic NTCP model which accounts for normal tissue recovery. This model shows NTCP is primarily affected by the recovery rate of the normal tissue as well as the fractional dose delivered to the OAR. In this prospective study, not all feathered organs received greater doses in the TFRT plans. This can be attributed to differences in the groups of individuals planning the non-TFRT IMRT plans and the TFRT plans. For future applications of TFRT, we propose using this dynamic NTCP model to predict the potential benefits of TFRT prior to delivery to steward resources. This would require further work to ascertain recovery rates for normal tissues. The acute toxicity observed in this study was as expected compared to our institutional standard.(6)

In this study, the five organs at risk most closely neighboring the high dose PTV were chosen to be feathered. Any OAR involved by tumor was not deliberately avoided or feathered so as to not compromise tumor control. Uninvolved submandibular glands were allowed to be feathered.(12) All normal organs permitted to be feathered were parallel organs. Parallel organs house functional subunits and therefore sparing partial volumes due to the spatial nature of temporally feathered radiation therapy has greater potential to decrease tissue toxicity as compared to serial organs. Additionally, to ensure safety, no serial structures were feathered. As part of this trial, whole organs were either deprioritized or spared. Future directions of TFRT can include the identification and selective sparing of functional subunits. For example the superior pharyngeal constrictors, excretory ducts of the salivary glands, and other partial volume structures implicated in preserving function could be avoided.(13,14) Notably, conventional dose volume histograms do not account for functional and structural heterogeneity and therefore cannot be used to assess the risk of inducing toxicity in this scenario. Lastly per protocol, five organs at risk were required to be feathered. These subplans were delivered in succession, and the time interval between the higher fractional doses was 7 days for each feathered organ. TFRT technique can be applied to any plan regardless of the number of organs feathered and the time between d_H_ and d_L_. With more understanding of normal tissue recovery, the number of organs feathered and resultant time between d_H_ and d_L_ may be titrated to the predicted NTCP profile.

Fifteen days were allowed between the time of CT simulation and commencement of radiation treatment in this phase I study, to test feasibility as the primary endpoint. The average time required for treatment planning was 10 days. Ongoing efforts to reduce the time required for treatment planning is important. Modern treatment planning systems which allow the creation of multiple can quicken the planning process. Additionally the use of multi-criteria optimization for IMRT planning could also allow for automation of the TFRT planning process. Future work will be focused on shortening the time required for TFRT planning. This will allow for larger subsequent clinical trials with well-defined clinical endpoints of toxicity. Lastly, TFRT may serve as a means of dose intensification to isotoxicity (same dose) in other disease sites where dose intensification is considered beneficial (e.g. pancreas).(15)

## Data Availability

All raw image data are available on request, otherwise they are included.

