## Supplemental Material for "*In Vivo* Assessment of the Safety of Standard Fractionation Temporally Feathered Radiation Therapy (TFRT) for Head and Neck Squamous Cell Carcinoma: An R-IDEAL Stage 1/2a First-in-Humans/Feasibility Demonstration of New Technology Implementation"

**Supplementary Appendix**

**Appendix A. Study Methods**

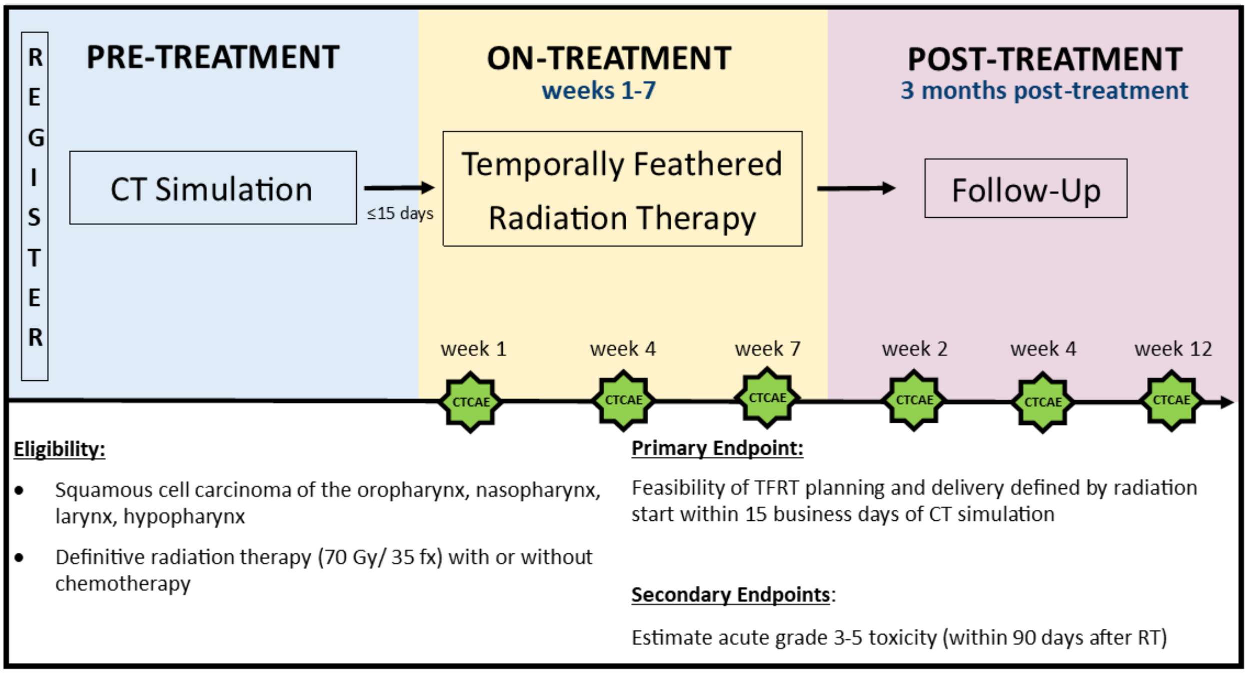


**Appendix B. TFRT planning techniques.**

| Compliance Criteria | | | |
|---|---|---|---|
| | **Per Protocol** | **Variation Acceptable** | **Deviation Unacceptable** |
| Total RT dose to PTV_7000 (to 95% of the PTV) | 70 Gy | None | none |
| Minimum dose ("cold spot" within PTV_7000, not including portion near (<8 mm skin) defined for a point that is 0.03 cc in size | 66.5 Gy (equals 95% prescribed dose) | <66.5 but >63 Gy | ≤63 Gy |
| Maximum dose ("hot spot" >1 cc) within PTV_7000 | ≤77 Gy | >77 Gy but ≤82 Gy | >82 Gy |
| Total RT dose to PTV_5600 (to 95% of the PTV) | 56 Gy | ≥45 but <56 Gy | <45 Gy |

| | | | |
|---|---|---|---|
| Total RT dose to PTV_6300 (to 95% of the PTV) | 63 Gy<br>**Required when applicable** | ≥52 but <63 Gy | <52 Gy |
| Total RT dose to spinal cord PRV (0.03 cc) | ≤50 Gy | ≥50 Gy but ≤52 Gy | >52 Gy |

Planning dosimetric constraints:

All dose constraints to critical structures include the volume of the critical structure outside of the planning target volume. Standard dose constraints were adapted from RTOG 1016 protocol (NCT01302834).

Spinal cord: 0.03 cc of the PRV should not exceed ≥50 Gy. 0.03 cc of the spinal cord should not exceed ≥45 Gy.

Brainstem: 0.03 cc of the brainstem should not exceed 60 Gy. 0.03 cc of the PRV brainstem should not exceed 63 Gy.

Lips: Reduce dose as much as possible, with goal of mean dose <20 Gy.

Oral Cavity: Reduce dose as much as possible, with goal of mean dose <30 Gy for the uninvolved oral cavity. Hot spots >60 Gy should be avoided as possible within the uninvolved oral cavity.

Parotid Glands: Each parotid gland should be optimized separately, with a goal of mean dose <26 Gy.

Contralateral Submandibular Glands: If contralateral nodal level IB is not targeted, goal is to reduce mean contralateral submandibular to <39 Gy.

OARpharynx: Reduce the dose as much as possible with goal mean dose <45 Gy.

Esophagus: Reduce the dose as much as possible, with goal mean dose <30 Gy.

Supraglottis: Reduce the dose as much as possible, with goal mean dose <45 Gy.

Larynx: Reduce the dose as much as possible, with goal mean dose <45 Gy.

GSL: Reduce the dose as much as possible, with goal mean dose <45 Gy.

**Appendix C. TFRT planning techniques.**

| **Therapist Treatment Delivery Timeout** |
|---|
| Two therapists must be present for the treatment timeout.<br><br>1. Current practices of verifying patient and treatment site must occur.<br>2. The treatment navigator in Mosaiq must be used to confirm the treatment plan fraction delivered the day before and determine the next appropriate fraction. Fractions will be |

| |
|---|
| delivered in a pattern of Plan A – Plan B – Plan C – Plan D – Plan E. For example, if plan B was delivered the day prior (i.e. on a Wednesday) then plan C must be delivered next for the next fraction (i.e. Thursday). In this example, Plan D would be delivered on Friday and Plan E would be delivered on Monday.<br>• In the event the patient misses a planned treatment fraction, the therapist must notify Eric Murray, CMD, Peng Qi, PhD, and Nikhil Joshi, MD. The determination can then be made in how to update the patient data in Mosaiq. Treatments should be resumed as soon as possible. The patient should resume therapy following the same pattern previously used A-B-C-D-E. Missed fractions should NOT be added to the end of the treatment schedule. Two fractions can never be delivered in the same day.<br>• No overrides are allowed to occur without the presence of a physicist. |

**Appendix D. Supplementary Dosimetric Data.**

**D.1 Dosimetric Data for Patient 1**

Organs feathered: oral cavity, right submandibular, left submandibular, OAR pharynx, supraglottis

| Target volume coverage for Patient 1 | | |
|---|---|---|
| Structure | Percent coverage achieved in non-TFRT IMRT plan | Dose achieved in composite TFRT plan |
| PTV_7000 | 95.22% receiving ≥70 Gy | 95.99% receiving ≥70 Gy |
| PTV_5600 | 95.35% receiving ≥56 Gy | 96.66% receiving ≥56 Gy |

| Dosimetric comparisons for Patient 1 | | |
|---|---|---|
| Structure | Dose (Gy) in non-TFRT IMRT Plan | Dose (Gy) in TFRT Plan |
| Maximum dose to 1 cc of PTV_7000 | 74.59 | 75.2 |
| Max point dose to brainstem | 12.20 | 12.03 |
| Max point dose to Brainstem PRV | 13.2 | 15.24 |
| Max point dose to Spinal Cord | 28.78 | 28.35 |
| Max point dose to Spinal Cord PRV | 34.8 | 34 |
| Mean dose to lips | 5.89 | 6.32 |
| Mean dose to oral cavity | 22.71 | 21.09 |
| Mean dose to right parotid | 12.9 | 12.03 |

| | | |
|---|---|---|
| Mean dose to left parotid | 22.26 | 22.35 |
| Mean dose to right submandibular gland | 40.44 | 37.16 |
| Mean dose to left submandibular gland | 42.76 | 43.5 |
| Mean dose to OAR pharynx | 40.85 | 43.78 |
| Mean dose to esophagus | 14.33 | 14.8 |
| Mean dose supraglottis | 33.32 | 30.85 |
| Mean dose to larynx | 15.78 | 18.39 |

*max point dose defined to 0.03 cc volume

| Fractional dose delivered to feathered organs in each subplan for patient 1 | | | | | |
|---|---|---|---|---|---|
| Treatment plan | Fractional dose to oral cavity (cGy) | Fractional dose to right submandibular (cGy) | Fractional dose left submandibular (cGy) | Fractional dose to supraglottis (cGy) | Fractional dose to OAR pharynx (cGy) |
| Subplan A | 79 | 94 | 115 | 84 | 118 |
| Subplan B | 57 | 37 | 113 | 84 | 124 |
| Subplan C | 56 | 99 | 153 | 85 | 123 |
| Subplan D | 55 | 101 | 120 | 105 | 123 |
| Subplan E | 55 | 100 | 120 | 86 | 136 |

**D.2 Dosimetric Data for Patient 2**

Organs feathered: oral cavity, right parotid, left submandibular, OAR pharynx, supraglottis

| Target volume coverage for Patient 2 | | |
|---|---|---|
| Structure | Percent coverage achieved in non-TFRT IMRT plan | Dose achieved in composite TFRT plan |
| PTV_7000 | 95.12% receiving ≥70 Gy | 95.48% receiving ≥70 Gy |
| PTV_5600 | 95.08% receiving ≥56 Gy | 95.74% receiving ≥56 Gy |

| Dosimetric comparisons for Patient 2 |
|---|

| Structure | Dose (Gy) in non-TFRT IMRT Plan | Dose (Gy) in TFRT Plan |
|---|---|---|
| Maximum dose to 1 cc of PTV_7000 | 75.43 | 74.95 |
| Max point dose to brainstem | 8.85 | 8.17 |
| Max point dose to Brainstem PRV | 10.51 | 11.03 |
| Max point dose to Spinal Cord | 26.58 | 21.83 |
| Max point dose to Spinal Cord PRV | 32.03 | 29.40 |
| Mean dose to lips | 4.16 | 4.72 |
| Mean dose to oral cavity | 22.83 | 21.41 |
| Mean dose to right parotid | 14.48 | 11.97 |
| Mean dose to left parotid | 8.76 | 8.70 |
| Mean dose to right submandibular gland | 62.47 | 64.51 |
| Mean dose to left submandibular gland | 30.66 | 27.81 |
| Mean dose to OAR pharynx | 37.13 | 37.54 |
| Mean dose to esophagus | 20.13 | 17.22 |
| Mean dose supraglottis | 34.23 | 36.73 |
| Mean dose to larynx | 20.27 | 19.44 |

*max point dose defined to 0.03 cc volume

| Fractional dose delivered to feathered organs in each subplan for patient 2 | | | | | |
|---|---|---|---|---|---|
| Treatment plan | Fractional dose to oral cavity (cGy) | Fractional dose to left submandibular (cGy) | Fractional dose right parotid (cGy) | Fractional dose to supraglottis (cGy) | Fractional dose to OAR pharynx (cGy) |
| Subplan A | 76 | 72 | 29 | 100 | 101 |
| Subplan B | 57 | 116 | 28 | 98 | 101 |

| | | | | | |
|---|---|---|---|---|---|
| Subplan C | 55 | 70 | 58 | 100 | 101 |
| Subplan D | 43 | 69 | 28 | 127 | 102 |
| Subplan E | 0.58 | 70 | 28 | 100 | 130 |

**D.3 Dosimetric Data for Patient 3**

Organs feathered: oral cavity, left parotid, right submandibular, OAR pharynx, supraglottis

| Target volume coverage for Patient 3 | | |
|---|---|---|
| Structure | Percent coverage achieved in non-TFRT IMRT plan | Dose achieved in composite TFRT plan |
| PTV_7000 | 95.50% receiving ≥70 Gy | 95.62% receiving ≥70 Gy |
| PTV_5600 | 95.93% receiving ≥56 Gy | 95.74% receiving ≥56 Gy |

| Dosimetric comparisons for Patient 3 | | |
|---|---|---|
| Structure | Dose (Gy) in non-TFRT IMRT Plan | Dose (Gy) in TFRT Plan |
| Maximum dose to 1 cc of PTV_7000 | 75.20 | 74.47 |
| Max point dose to brainstem | 17.81 | 20.97 |
| Max point dose to Brainstem PRV | 22.40 | 26.68 |
| Max point dose to Spinal Cord | 24.96 | 24.65 |
| Max point dose to Spinal Cord PRV | 34.80 | 38.93 |
| Mean dose to lips | 7.10 | 5.63 |
| Mean dose to oral cavity | 32.91 | 27.3 |
| Mean dose to right parotid | 22.73 | 21.2 |
| Mean dose to left parotid | 31.80 | 30.67 |
| Mean dose to right submandibular gland | 38.57 | 36.80 |

| | | |
|---|---|---|
| Mean dose to left submandibular gland | 65.39 | 65.63 |
| Mean dose to OAR pharynx | 46.08 | 44.46 |
| Mean dose to esophagus | 14.91 | 10.56 |
| Mean dose supraglottis | 26.30 | 26.33 |
| Mean dose to larynx | 19.21 | 19.54 |

*max point dose defined to 0.03 cc volume

| Fractional dose delivered to feathered organs in each subplan for patient 3 | | | | | |
|---|---|---|---|---|---|
| Treatment plan | Fractional dose to oral cavity (cGy) | Fractional dose to right submandibular (cGy) | Fractional dose left parotid (cGy) | Fractional dose to supraglottis (cGy) | Fractional dose to OAR pharynx (cGy) |
| Subplan A | 91 | 99 | 83 | 69 | 125 |
| Subplan B | 76 | 124 | 84 | 70 | 125 |
| Subplan C | 74 | 101 | 104 | 70 | 126 |
| Subplan D | 74 | 101 | 84 | 96 | 126 |
| Subplan E | 74 | 100 | 84 | 72 | 133 |

**D.4 Dosimetric Data for Patient 4**

Organs feathered: oral cavity, left parotid, right submandibular, OAR pharynx, supraglottis

| Target volume coverage for Patient 4 | | |
|---|---|---|
| Structure | Percent coverage achieved in non-TFRT IMRT plan | Dose achieved in composite TFRT plan |
| PTV_7000 | 95.31% receiving ≥70 Gy | 95.30% receiving ≥70 Gy |
| PTV_5600 | 95.37% receiving ≥56 Gy | 96.63% receiving ≥56 Gy |

| Dosimetric comparisons for Patient 4 | | |
|---|---|---|
| Structure | Dose (Gy) in non-TFRT IMRT Plan | Dose (Gy) in TFRT Plan |

| | | |
|---|---|---|
| Maximum dose to 1 cc of PTV_7000 | 75.06 | 75.8 |
| Max point dose to brainstem | 15.93 | 22.82 |
| Max point dose to Brainstem PRV | 20.25 | 27.61 |
| Max point dose to Spinal Cord | 25.73 | 24.61 |
| Max point dose to Spinal Cord PRV | 30.57 | 33.58 |
| Mean dose to lips | 7.82 | 8.98 |
| Mean dose to oral cavity | 54.77 | 54.99 |
| Mean dose to right parotid | 25.94 | 20.92 |
| Mean dose to left parotid | 31.32 | 31.34 |
| Mean dose to right submandibular gland | 61.64 | 56.70 |
| Mean dose to left submandibular gland | 68.07 | 68.86 |
| Mean dose to OAR pharynx | 47.02 | 44.98 |
| Mean dose to esophagus | 18.48 | 17.87 |
| Mean dose supraglottis | 46.63 | 44.99 |
| Mean dose to larynx | 17.21 | 18.54 |

*max point dose defined to 0.03 cc volume

| Fractional dose delivered to feathered organs in each subplan for patient 4 | | | | | |
|---|---|---|---|---|---|
| Treatment plan | Fractional dose to right submandibular (cGy) | Fractional dose to left partoid (cGy) | Fractional dose supraglottis (cGy) | Fractional dose to oral cavity (cGy) | Fractional dose to OAR pharynx (cGy) |
| Subplan A | 181 | 85 | 128 | 156 | 127 |
| Subplan B | 156 | 106 | 128 | 157 | 127 |
| Subplan C | 156 | 87 | 132 | 157 | 126 |

| | | | | | |
|---|---|---|---|---|---|
| Subplan D | 156 | 86 | 128 | 160 | 127 |
| Subplan E | 161 | 85 | 127 | 156 | 136 |
